# Susceptibility to postmortem (co)-pathologies in antemortem atrophy-based subtypes of Alzheimer’s disease

**DOI:** 10.1101/2021.09.06.21263162

**Authors:** Rosaleena Mohanty, Daniel Ferreira, Simon Frerich, J-Sebastian Muehlboeck, Michel Grothe, Eric Westman, for the Alzheimer’s Disease Neuroimaging Initiative

## Abstract

**Objectives:** To investigate whether antemortem atrophy-based subtypes of Alzheimer’s disease (AD) may be differentially susceptible to individual or concomitance of AD and non-AD (co)-pathologies, assessed neuropathologically at postmortem.

**Methods:** We selected 31 individuals from the AD neuroimaging initiative with: an antemortem magnetic resonance imaging scan evaluating brain atrophy available within two years before death; an antemortem diagnosis of AD dementia or prodromal AD; and postmortem neuropathological confirmation of AD. Antemortem atrophy-based subtypes was modeled as a continuous phenomenon in terms of two recently proposed dimensions: *typicality* (ranging from *limbic-predominant AD* to *hippocampal-sparing AD* subtypes) and *severity* (ranging from *typical AD* to *minimal atrophy AD* subtypes). Postmortem neuropathological evaluation included global and regional outcomes: AD hallmark pathologies of amyloid-beta and tau; non-AD co-pathologies of alpha-synuclein Lewy body and TDP-43; and the overall concomitance across these four (co)-pathologies. Partial correlation and linear regression models were used to assess the association between antemortem atrophy-based subtypes and postmortem neuropathological outcomes.

**Results:** We observed significant global and regional associations between antemortem typicality and postmortem (co)-pathologies including tau, alpha-synuclein Lewy bodies and TDP-43. Antemortem typicality demonstrated stronger regional associations with concomitance of multiple postmortem (co)-pathologies in comparison to antemortem severity. Our findings suggest the following susceptibilities of atrophy-based subtypes: limbic-predominant AD towards higher burden of tau and TDP-43 pathologies while hippocampal-sparing AD towards lower burdens; limbic-predominant AD and typical AD towards higher burden of alpha-synuclein Lewy body pathology while hippocampal-sparing AD and minimal-atrophy AD towards lower burdens.

**Discussion:** Through a direct antemortem-to-postmortem validation, our study highlights the importance of understanding heterogeneity in AD in relation to concomitance of AD and non-AD pathologies. Our findings provide a deeper understanding of both global and regional vulnerabilities of the biological subtypes of AD brain towards (co)-pathologies. Relative involvement of both AD hallmark and non-AD (co)-pathologies will enhance prevailing knowledge of biological heterogeneity in AD and could thus, contribute towards tracking disease progression and designing clinical trials in the future.

## Introduction

“Pure” Alzheimer’s disease (AD), epitomized by the hallmarks of amyloid beta (Aβ) and tau neurofibrillary tangles (NFT), is increasingly recognized as not being the most prevalent form of the disease^1–3^. Concomitant forms of pathological proteins such as α-synuclein (α-syn) and TAR DNA-binding protein 43 (TDP-43) have been reported in over 40%^4^ and 50%^5^ of the AD cases respectively.

Does this multimorbid view of AD brain suggest that brain atrophy may be downstream to not only the AD hallmark pathologies, but also to the interactions with one or more concomitant pathologies? De Flores et al. examined medial temporal atrophy and reported that postmortem tau pathology was associated with antemortem posterior hippocampal atrophy, and postmortem TDP-43 pathology was associated with antemortem anterior medial temporal atrophy^6^. Medial temporal atrophy, although a common characteristic, is not always observed in AD. Converging evidence suggests that biological heterogeneity in AD may manifest as distinct atrophy subtypes: *typical* AD, *limbic-predominant* AD, *hippocampal-sparing* AD, and *minimal-atrophy* AD^7^, with the last two not showing medial temporal atrophy. Thus, revising the initial question, we ask: does this multimorbid view of AD brain suggest that brain atrophy *subtypes* may be downstream to not only the AD hallmark pathologies, but also to the interactions with one or more concomitant pathologies? To our knowledge, the answer to this question is yet to be explored.

We currently lack *in vivo* biomarkers to assess pathologies such as α-syn and TDP-43. Therefore, we set out to investigate the relationship between antemortem atrophy subtypes and postmortem neuropathological profiles in AD, in the first study of this kind to our knowledge. Despite limited availability of postmortem data, recent studies have shown added value and face validity of studying them^8,9^, given that postmortem assessments serve as the current gold standard in AD. Therefore, we herein pose our key research questions: (a) are antemortem atrophy subtypes of AD related to individual and/or concomitance of (co)-pathologies at postmortem? (b) does this subtype-to-(co)-pathology relationship vary by brain region? To this end, we modeled antemortem atrophy subtypes in AD, as assessed by magnetic resonance imaging (MRI). We operationalized AD subtypes as a continuous phenomenon^10^, following the recent conceptual framework for AD subtypes^7^, and examined the direct relationship to postmortem global and regional (co)-pathologies (Aβ, tau, α-syn Lewy body, TDP-43) at group-and individual-levels.

## Methods

### Participants

Participants were selected from the Alzheimer’s Disease Neuroimaging Initiative (ADNI) database (PI: M. Weiner; http://adni.loni.usc.edu/). Launched in 2003, the goal of the ADNI is to test and use biomarkers, clinical and neuropsychological assessments to track disease progression in AD. We included data from participants who had antemortem MRI and postmortem neuropathological assessments (Version 11, 04/12/2018). **Figure 1** shows the selection criteria for this study. Our final cohort comprised 31 participants such that the antemortem MRI was available within 2 years prior to death (to avoid long antemortem-to-postmortem interval being a potential confound) and neuropathological assessment showed intermediate or high AD neuropathologic change (i.e., pathology-confirmed AD dementia)^11^. All the ADNI protocols were approved by the institutional review boards of each participating institution. All participants provided written and informed consent in accordance with the Declaration of Helsinki.

**Figure 1.**
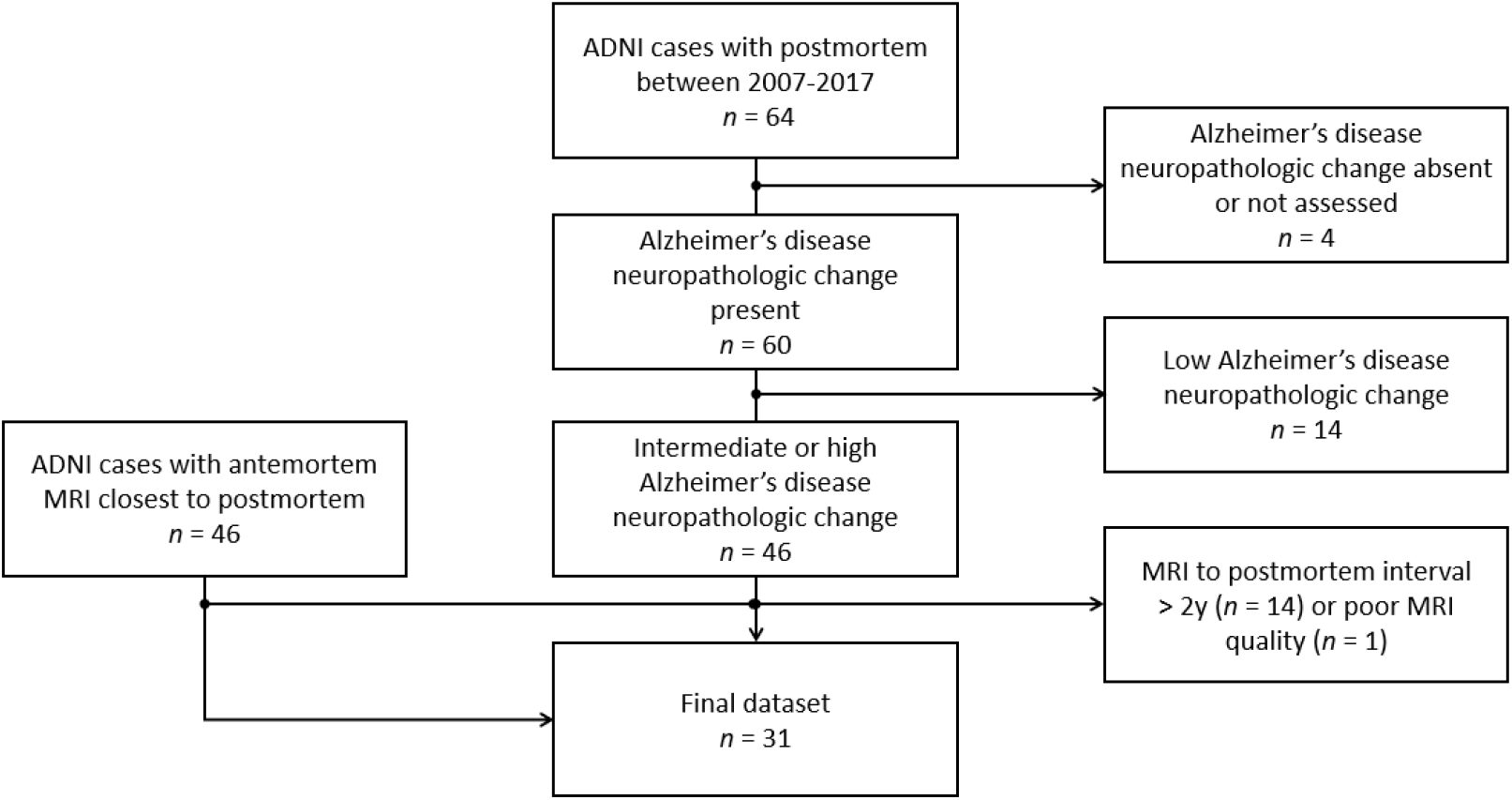
Flowchart of selection of study cohort ADNI=Alzheimer’s disease neuroimaging initiative; MRI=magnetic resonance imaging.

### Antemortem neuroimaging and cognition

MRI scans were acquired on 1.5T or 3T scanners with T1-weighted sagittal 3D magnetization-prepared rapid gradient echo (MPRAGE) sequences (detailed ADNI imaging protocols: adni.loni.usc.edu/methods/). MRI were processed cross-sectionally using FreeSurfer 6.0.0 (http://freesurfer.net/), automated through TheHiveDB system^12^. Resulting segmentations were visually screened for quality control. Screened scans were included for subsequent analyses. Automatic region of interest parcellation yielded volume of 34 cortical areas^13^ and 7 subcortical areas^14^ per hemisphere, serving as a measure of brain atrophy. We used mini mental examination (MMSE)^15^ corresponding to the MRI visit as the main outcome to evaluate the level of global cognitive impairment.

### Antemortem atrophy subtypes

Following the recently proposed conceptual framework for AD subtypes^7^, we quantified MRI-based atrophy subtypes in terms of two principal dimensions: typicality and severity. Given the limited sample size, we modeled atrophy subtypes on a continuous scale for greater sensitivity^16^ rather than categorizing individuals into subgroups/categorical subtypes. *Typicality* was proxied by the ratio of hippocampal volume to whole cortical volume (referred to as *H:C* hereon), similar to the index adopted by the original neuropathological subtyping study^17^. *Severity* was proxied by the global brain atrophy index, measured by the ratio of whole brain volume to volume of cerebrospinal fluid^18^ (referred to as *BV:CSF* hereon), such that lower values of the index correspond to more atrophy and hence, higher severity.

### Postmortem Neuropathological Assessment

Neuropathological assessments were conducted as part of the ADNI neuropathology core (neuropathologist: Dr. Nigel Cairns, the Knight Alzheimer’s Disease Research Center, Washington University School of Medicine, St. Louis, http://adni.loni.usc.edu/about/#core-container)^19^. Assessments followed the NIA-AA guidelines for the neuropathologic assessment of AD^11^ (https://www.alz.washington.edu/NONMEMBER/NP/npguide10.pdf).

To investigate our first research question of whether antemortem atrophy subtypes of AD may be related to individual postmortem (co)-pathologies, we examined *global (co)-pathological outcomes:* (a) AD-specific neuropathological measures (semi-quantitative) of the AD neuropathologic change (ADNC), comprising the Thal phase of regional distribution of Aβ plaques (A0-A3), the Braak stage of tau NFT (B0-B3), and the Consortium to Establish a Registry for Alzheimer’s Disease (CERAD) scores for density of neuritic plaques (C0-C3)^11^; and (b) non-AD-specific neuropathological measures (binarized for presence/absence) including Lewy body (LB) pathology, assessed in the brainstem, limbic region, neocortex, amygdala and olfactory bulb as per the modified McKeith criteria^11,20^, and TDP-43 pathology assessed as immunoreactive inclusions in the spinal cord, amygdala, hippocampus, entorhinal cortex/inferior temporal gyrus, and frontal neocortex^21^.

To investigate our second research question of whether antemortem atrophy subtypes of AD may be related to postmortem (co)-pathologies varying by brain regions, we examined *regional (co)-pathological outcomes:* we analyzed brain regions most relevant to atrophy subtypes in AD^7^, i.e., structures of the medial temporal lobe including the hippocampus at the level of lateral geniculate nucleus (CA1, dentate gyrus, parahippocampal gyrus), amygdala and entorhinal cortex, and structures of the association cortex including the middle frontal gyrus, superior and middle temporal gyri and inferior parietal lobe (angular gyrus). We focused on: (a) AD-specific neuropathological measures (semi-quantitative) of Aβ (diffuse plaque, DP; cored plaque, CP; cerebral amyloid angiopathy, CAA) and tau (NFT; neuritic plaque, NP; glial); and (b) non-AD-specific neuropathological measures (semi-quantitative) of α-syn (LB) and TDP-43 (neuronal cytoplasmic inclusion, NCI; dystrophic neurite, DN). Forms of the investigated proteins showing little to no variability across individuals and/or regions were excluded from analyses (e.g., α-syn as glial cytoplasmic inclusion, GCI; TDP-43 as neuronal intranuclear inclusions, NII; etc.).

To investigate our research questions of whether antemortem atrophy subtypes of AD may be related to concomitance of postmortem (co)-pathologies and may vary regionally, we evaluated the *total number of pathologies* present per region as an outcome: each pathology was binarized (present/absent) and summed, considering all forms of the aforementioned AD-specific and non-AD-specific pathologies (ranging from 0 to 9).

### Statistical analysis

We analyzed the association between antemortem atrophy subtypes (typicality and severity as continuous independent variables in separate models) and cognition as well as postmortem (co)-pathologies (dependent variable) using partial correlations for semi-quantitative outcomes (global and regional measures) and linear regression for continuous outcomes (total number of pathologies i.e., concomitance). All models were controlled for age at MRI scan and MRI scanner field strength. Given that postmortem neuropathological data are rare, we report significant results at uncorrected *p*-value < 0.05, akin to previous studies utilizing postmortem data22,23.

All statistical analyses and visualizations were conducted using MATLAB R2020b (The MathWorks, Inc., Natick, Massachusetts, USA).

## Results

### Participants

**Table 1** shows the demographic and ante-/post-mortem clinical profiles of the cohort. The age at antemortem MRI was 80 ± 6.7 y while the age at death was 81.2 ± 6.7 y. The level of global cognitive impairment antemortem was 18.2 ± 6.7 on MMSE.

**Table 1.**
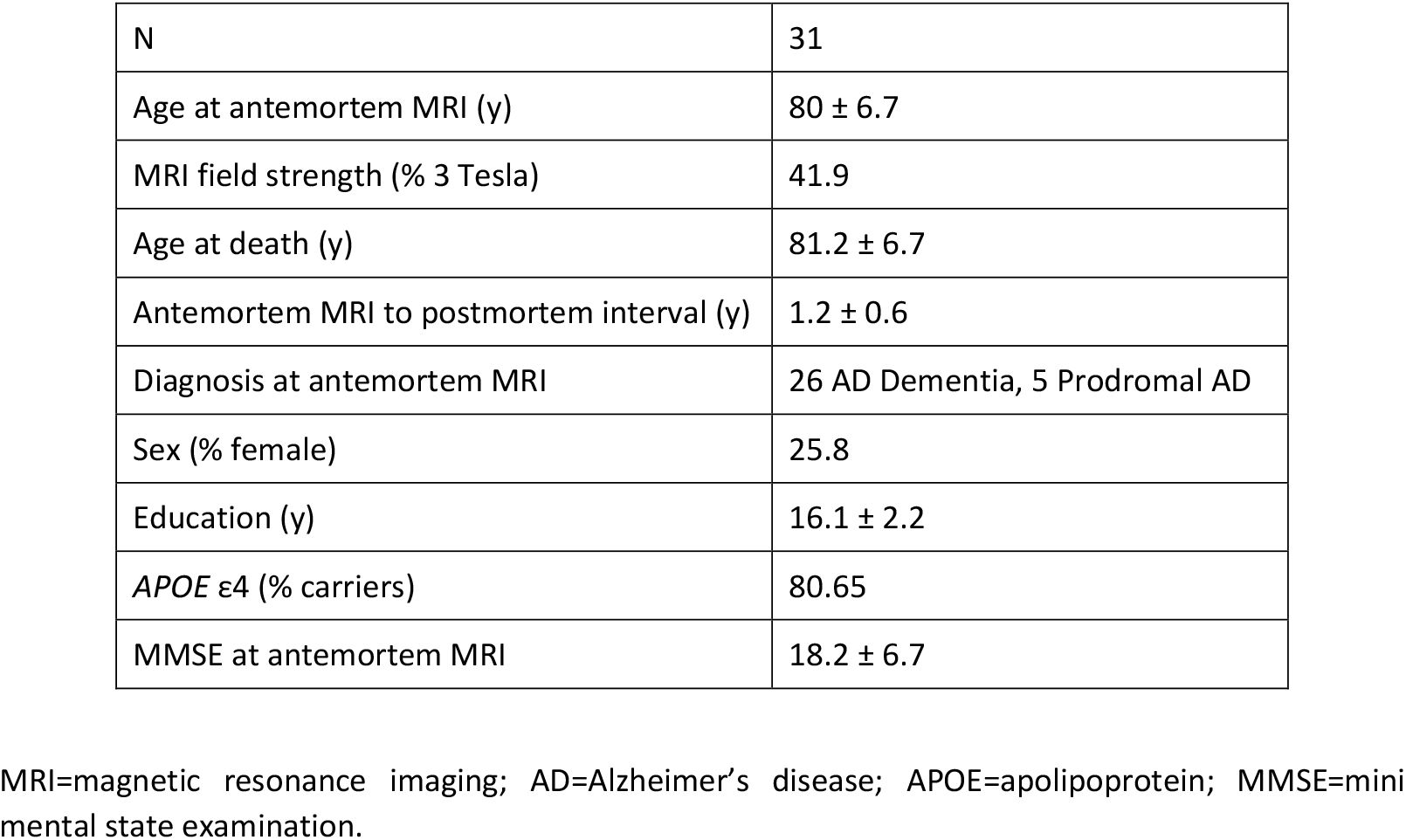
Characteristics of the selected cohort

|  |  |
| --- | --- |
| N | 31 |
| Age at antemortem MRI (y) | 80 ± 6.7 |
| MRI field strength (% 3 Tesla) | 41.9 |
| Age at death (y) | 81.2 ± 6.7 |
| Antemortem MRI to postmortem interval (y) | 1.2 ± 0.6 |
| Diagnosis at antemortem MRI | 26 AD Dementia, 5 Prodromal AD |
| Sex (% female) | 25.8 |
| Education (y) | 16.1 ± 2.2 |
| *APOE* ε4 (% carriers) | 80.65 |
| MMSE at antemortem MRI | 18.2 ± 6.7 |

MRI=magnetic resonance imaging; AD=Alzheimer's disease; APOE=apolipoprotein; MMSE=mini mental state examination.

### Antemortem atrophy subtypes

**Figure 2A** shows the atrophy subtypes in antemortem MRI, characterized by the continuous-scale measures of typicality (H:C) and severity (BV:CSF). We show four exemplars to illustrate the extremes on each scale. On the typicality scale, case RID 1203 represents hippocampal-sparing AD towards the higher extreme while case RID 1393 represents limbic-predominant AD towards the lower extreme. Similarly, on the severity scale, case RID 1271 represents typical AD towards the lower extreme (higher severity) while case RID 1425 represents minimal-atrophy AD towards the higher extreme (lower severity). The association between typicality and severity was not statistically significant (r=0.31, *p*=0.09). Antemortem severity (r=0.51, *p*=0.042) but not typicality (r=0.34, *p*=0.34) was significantly associated with antemortem MMSE.

**Figure 2.**
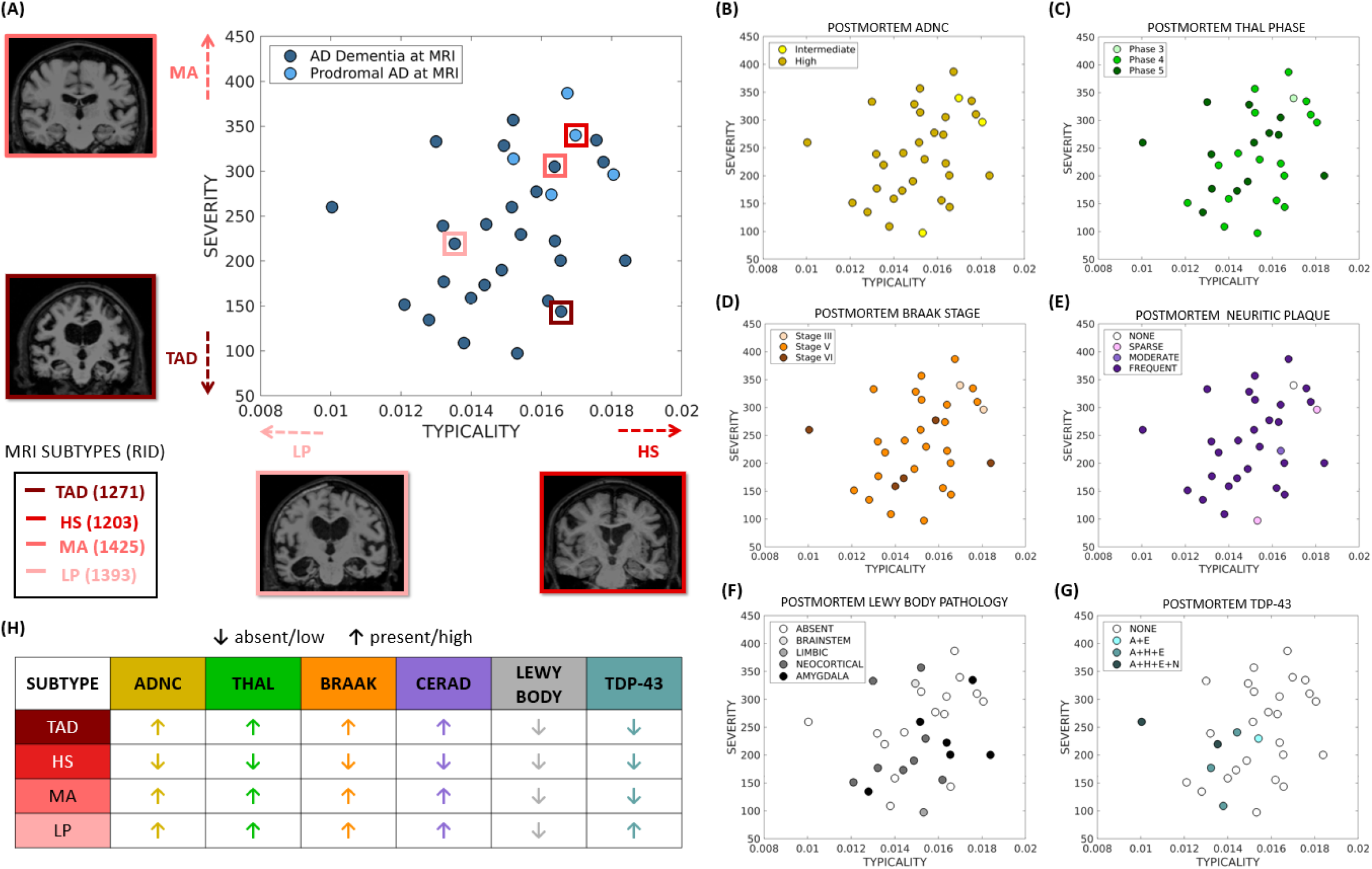
Distribution of (**A**) antemortem MRI-based heterogeneity, (**B-G**) postmortem neuropathology superposed on MRI-based heterogeneity, and (**H**) summary of postmortem neuropathology in four case studies AD=Alzheimer’s disease; MRI=magnetic resonance imaging; TAD=typical AD; HS=hippocampal-sparing AD; MA=minimal-atrophy AD; LP=limbic-predominant AD; ADNC=AD neuropathological change; RID=Assigned individual ID in the AD Neuroimaging Initiative dataset; TDP=TAR DNA-binding protein; A+E=TDP-43 present in the amygdala and entorhinal/inferior temporal cortex; A+H+E=TDP-43 immunoreactive inclusions are present in the amygdala, hippocampus and entorhinal/inferior temporal cortex; A+H+E+N=TDP-43 immunoreactive inclusions are present in the amygdala, hippocampus, entorhinal/inferior temporal cortex and neocortex; CERAD=Consortium to Establish a Registry for Alzheimer’s Disease score to assess neuritic plaques. All plots show antemortem MRI-based typicality on the horizontal scale, proxied by the 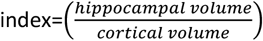; All plots show antemortem MRI-based severity on the vertical scale, proxied by the global brain atrophy 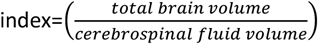 (higher values correspond to lower severity).

### Association between antemortem typicality and postmortem (co)-pathologies

**Table 2** shows the association between typicality and global neuropathological measures. Typicality was significantly associated with Thal Aβ phase (96.8% at A3, i.e., Phase 4-5; **Figure 2C**), neuritic plaques (87.1% at C3, i.e., frequent neuritic plaques; **Figure 2E**) and TDP-43 inclusions in each of amygdala, hippocampus, entorhinal cortex, and neocortex (**Figure 2G**). These associations were negative, i.e., higher/lower value of H:C (hippocampal-sparing/limbic predominant AD) was associated with lower/higher burden of (co)-pathology.

**Table 2.** Association between antemortem atrophy and global neuropathological measures

| Postmortem pathology | Antemortem Typicality<br>rho (*p*) | Antemortem Severity<br>rho (*p*) |
| --- | --- | --- |
| Thal Aβ phase | **-0.37 (0.039)** | 0.03 (0.87) |
| Braak Tau stage | -0.26 (0.15) | -0.26 (0.18) |
| Neuritic plaque | **-0.36 (0.047)** | 0.04 (0.84) |
| **Lewy body pathology** |  |  |
| Brainstem-predominant | -0.042 (0.83) | 0.28 (0.13) |
| Limbic | -0.002 (0.99) | **-0.38 (0.04)** |
| Neocortical | -0.35 (0.06) | -0.15 (0.45) |
| Amygdala-predominant | 0.24 (0.20) | -0.10 (0.59) |
| **TDP-43 inclusions** |  |  |
| Amygdala | **-0.53 (0.004)** | -0.28 (0.16) |
| Hippocampus | **-0.48 (0.012)** | -0.15 (0.46) |
| Entorhinal cortex/inferior temporal cortex | **-0.42 (0.029)** | -0.15 (0.44) |
| Neocortex | **-0.39 (0.041)** | -0.02 (0.91) |

TDP=TAR DNA-binding protein; Associations between typicality or severity and individual pathologies were evaluated using partial correlation, adjusted for field strength and age at scan; rho=Spearman partial correlation coefficient; *p* < 0.05 are shown in **bold**.

**Table 3** shows the association between typicality and regional neuropathological measures. Typicality was significantly associated with: (a) p-Tau (NFT) in the dentate gyrus; (b) p-α-Syn LB in the parahippocampal gyrus and superior/middle temporal gyri; (c) p-TDP-43 (NCI) in the parahippocampal gyrus, dentate gyrus, entorhinal cortex, amygdala, superior/middle temporal gyri; and (d) p-TDP-43 (DN) in the entorhinal cortex. These associations were negative, i.e., higher/lower value of H:C (hippocampal-sparing/limbic-predominant AD) was associated with lower/higher burden of (co)-pathology. Further, typicality was positively associated with the number of (co)-pathologies in the parahippocampal gyrus, dentate gyrus, entorhinal cortex, amygdala and superior/middle temporal gyri. This suggests that higher/lower value of H:C (hippocampal-sparing/limbic-predominant AD) showed higher/lower concomitance of multiple (co)-pathologies (**Figure 3**). None of the cases demonstrated presence of all nine forms of (co)-pathologies.

**Table 3.** Association between antemortem MRI-based heterogeneity and regional neuropathological measures

| Antemortem typicality versus postmortem neuropathology |  |  |  |  |  |  |  |  |
| --- | --- | --- | --- | --- | --- | --- | --- | --- |
|  | PHG | DG | CA1 | ERC | AMYG | IPL | STG | MFG |
| Aβ (DP) | - | -0.06<br>(0.74) | -0.21<br>(0.28) | - | - | - | - | - |
| Aβ (CP) | -0.21<br>(0.28) | -0.07<br>(0.71) | -0.05<br>(0.78) | -0.21<br>(0.29) | 0.002<br>(0.99) | - | - | - |
| Aβ (CAA) | -0.22<br>(0.24) | -0.31<br>(0.11) | -0.28<br>(0.13) | -0.32<br>(0.1) | -0.18<br>(0.37) | -0.01<br>(0.96) | -0.12<br>(0.53) | -0.03<br>(0.89) |
| p-Tau (NFT) | - | **-0.45**<br>**(0.014)** | - | - | - | -0.15<br>(0.43) | - | -0.13<br>(0.49) |
| p-Tau (NP) | -0.11<br>(0.56) | -0.13<br>(0.49) | 0.12<br>(0.54) | -0.21<br>(0.29) | -0.09<br>(0.64) | -0.36<br>(0.053)* | -0.37<br>(0.054)* | -0.26<br>(0.18) |
| p-Tau (Glial) | -0.13<br>(0.51) | -0.34<br>(0.07) | 0.29<br>(0.12) | -0.12<br>(0.55) | -0.36<br>(0.06) | -0.19<br>(0.32) | -0.19<br>(0.31) | -0.08<br>(0.68) |
| p-α-Syn LB | **-0.44**<br>**(0.017)** | - | -0.10<br>(0.59) | -0.21<br>(0.28) | -0.09<br>(0.66) | 0.07<br>(0.69) | **-0.38**<br>**(0.046)** | -0.14<br>(0.48) |
| p-TDP-43 (NCI) | **-0.43**<br>**(0.023)** | **-0.41**<br>**(0.029)** | -0.25<br>(0.2) | **-0.43**<br>**(0.024)** | **-0.54**<br>**(0.003)** | -0.20<br>(0.3) | **-0.38**<br>**(0.048)** | -0.17<br>(0.39) |
| p-TDP-43 (DN) | -0.25<br>(0.21) | - | -0.20<br>(0.3) | **-0.39**<br>**(0.041)** | -0.12<br>(0.56) | -0.20<br>(0.3) | -0.34<br>(0.09) | - |
| Total number of pathologies | **0.29**<br>**(0.002)** | **0.46**<br>**(0.0002)** | 0.07<br>(0.28) | **0.25**<br>**(0.01)** | **0.17**<br>**(0.04)** | 0.22<br>(0.08) | **0.23**<br>**(0.02)** | 0.06<br>(0.3) |
| Antemortem severity versus postmortem neuropathology |  |  |  |  |  |  |  |  |
|  | PHG | DG | CA1 | ERC | AMYG | IPL | STG | MFG |
| Aβ (DP) | - | 0.24<br>(0.21) | -0.2<br>(0.31) | - | - | - | - | - |
| Aβ (CP) | -0.2<br>(0.31) | -0.09<br>(0.64) | -0.05<br>(0.8) | -0.23<br>(0.23) | -0.08<br>(0.69) | - | - | - |
| Aβ (CAA) | 0.22<br>(0.25) | 0.15<br>(0.44) | 0.06<br>(0.76) | 0.18<br>(0.35) | 0.02<br>(0.93) | 0.18<br>(0.34) | 0.04<br>(0.84) | -0.19<br>(0.31) |
| p-Tau (NFT) | - | -0.33<br>(0.08) | - | - | - | 0.14<br>(0.48) | - | -0.12<br>(0.55) |
| p-Tau (NP) | -0.33<br>(0.07) | 0.06<br>(0.74) | 0.04<br>(0.84) | -0.23<br>(0.23) | -0.08<br>(0.67) | 0.05<br>(0.78) | -0.25<br>(0.19) | -0.19<br>(0.32) |
| p-Tau (Glial) | -0.01<br>(0.96) | -0.005<br>(0.98) | 0.15<br>(0.43) | 0.11<br>(0.56) | -0.28<br>(0.14) | -0.19<br>(0.33) | 0.07<br>(0.71) | -0.10<br>(0.59) |
| p-α-Syn LB | -0.23<br>(0.23) | - | 0.01<br>(0.98) | -0.37<br>(0.052)* | -0.26<br>(0.17) | 0.05<br>(0.79) | -0.19<br>(0.34) | -0.02<br>(0.93) |
| p-TDP-43 (NCI) | -0.18<br>(0.35) | -0.03<br>(0.89) | -0.22<br>(0.28) | -0.17<br>(0.4) | -0.28<br>(0.15) | -0.06<br>(0.75) | -0.05<br>(0.82) | -0.33<br>(0.08) |
| **p-TDP-43 (DN)** | -0.03<br>(0.85) | - | -0.06<br>(0.75) | -0.02<br>(0.92) | 0.02<br>(0.91) | -0.06<br>(0.75) | 0.01<br>(0.95) | - |
| **Total number of pathologies** | 0.04<br>(0.36) | 0.1<br>(0.92) | 0.03<br>(0.77) | 0.14<br>(0.1) | **0.18**<br>**(0.03)** | 0.13<br>(0.6) | 0.06<br>(0.72) | 0.09<br>(0.17) |

MRI=magnetic resonance imaging; PHG=hippocampus at the level of lateral geniculate nucleus including parahippocampal gyrus; DG=hippocampus at the level of lateral geniculate nucleus including dentate gyrus; CA1=hippocampus at the level of lateral geniculate nucleus including cornu ammonis1 subfield; ERC=entorhinal cortex; AMYG=amygdala; IPL=inferior parietal lobe (angular gyrus); STG=superior and middle temporal gyri; MFG=middle frontal gyrus; Aβ =beta-amyloid; DP=diffuse plaque; CP=cored plaque; CAA=cerebral amyloid angiopathy; p-Tau=phosphorylated tau; NFT=neurofibrillary tangle; NP=neuritic plaque; p-α-Syn=phosphorylated alpha-synuclein; LB=Lewy body; p-TDP-43=phosphorylated TAR DNA-binding protein; NCI=neuronal cytoplasmic inclusion; DN=dystrophic neurite; Associations between typicality or severity and individual pathologies were evaluated using partial correlation, adjusted for field strength and age at scan; Spearman partial correlation coefficient (*p*-values) are reported; Associations between typicality/severity and the total number of pathologies was evaluated using linear regression, adjusted for field strength and age at scan; Correlation coefficient (*p*-value) are reported; *p* < 0.05 are shown in **bold**; marginally significant associations are marked by \*.

**Figure 3.**
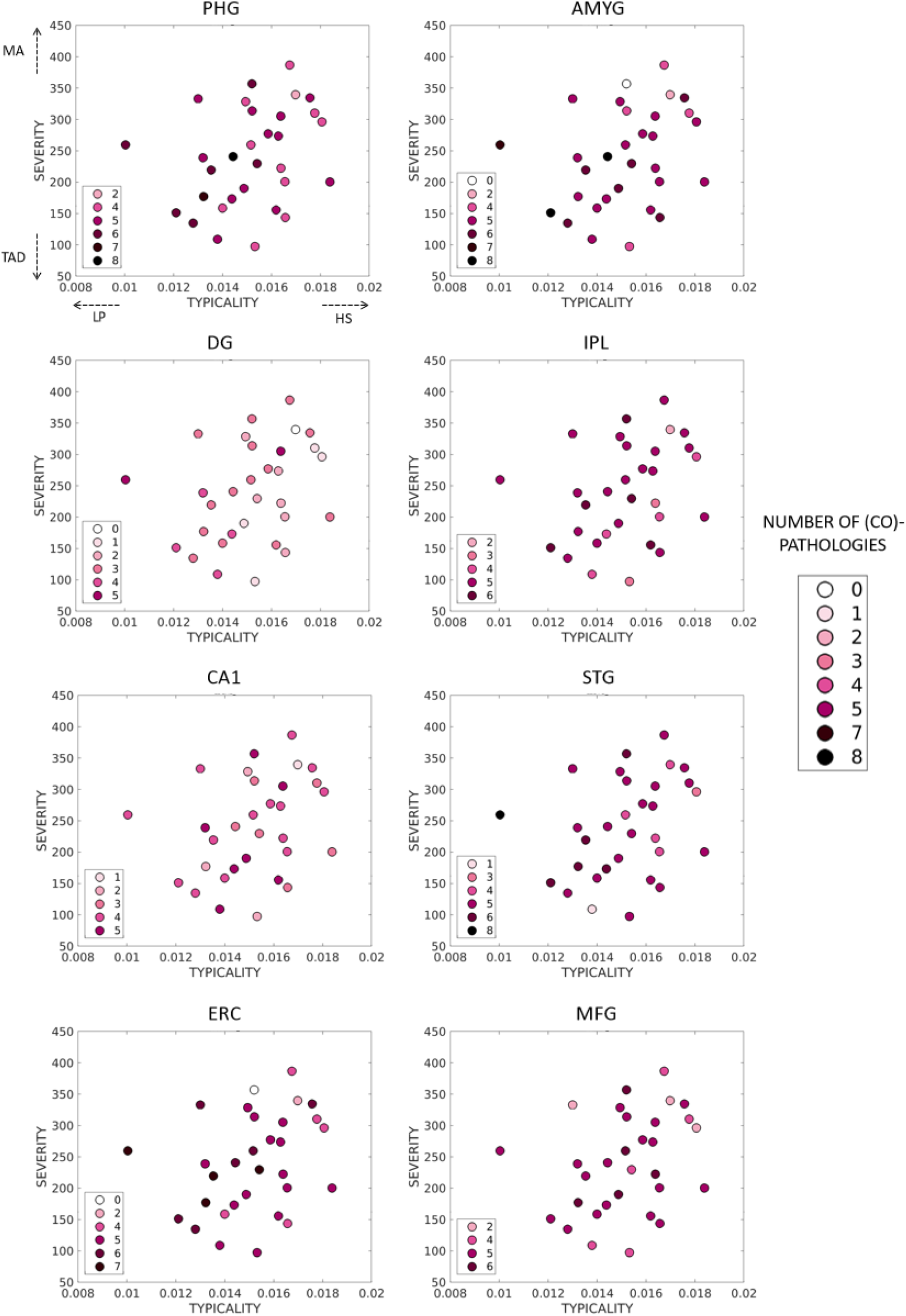
Regional distribution of postmortem (co)-pathologies superposed on antemortem MRI-based heterogeneity Number of (co)-pathologies was evaluated as the sum of presence of individual pathologies including Aβ (DP), Aβ (CP), Aβ (CAA), p-Tau (NFT), p-Tau (NP), p-Tau (Glial), p-α-Syn LB, p-TDP-43 (NCI), p-TDP-43 (DN); Aβ=beta-amyloid; DP=diffuse plaque; CP=cored plaque; CAA=cerebral amyloid angiopathy; p-Tau=phosphorylated tau; NFT=neurofibrillary tangle; NP=neuritic plaque; p-α-Syn=phosphorylated alpha-synuclein; LB=Lewy body; p-TDP-43=phosphorylated TAR DNA-binding protein; NCI=neuronal cytoplasmic inclusion; DN=dystrophic neurite; PHG=hippocampus at the level of lateral geniculate nucleus including parahippocampal gyrus; DG=hippocampus at the level of lateral geniculate nucleus including dentate gyrus; CA1=hippocampus at the level of lateral geniculate nucleus including cornu ammonis1 subfield; ERC=entorhinal cortex; AMYG=amygdala; IPL=inferior parietal lobe (angular gyrus); STG=superior and middle temporal gyri; MFG=middle frontal gyrus; TAD=typical AD; HS=hippocampal-sparing AD; MA=minimal-atrophy AD; LP=limbic-predominant AD.

### Association between antemortem severity and postmortem (co)-pathologies

**Table 2** shows the association between antemortem severity and global neuropathological measures. Severity was significantly associated with Lewy body pathology in the limbic regions (**Figure 2F**). This association was negative, i.e., higher/lower value of BV:CSF (minimal-atrophy/typical AD) was associated with lower/higher burden of Lewy body pathology in the region.

**Table 3** shows that there were no significant associations between antemortem severity and regional neuropathological measures. However, severity was positively associated with the number of (co)-pathologies in the amygdala. This indicates that higher/lower value of BV:CSF (minimal-atrophy/typical AD) showed higher/lower concomitance of multiple (co)-pathologies (**Figure 3**).

### Case studies

Below we present antemortem-to-postmortem profiles of four exemplars (**Figure 2A, H**): Case RID 1203 had prodromal AD antemortem, a higher H:C, relatively spared hippocampi on MRI and thus, presented as *hippocampal-sparing AD*. At postmortem, case RID 1203 had relatively lower burdens of Aβ and tau, however, were devoid of neuritic plaque, Lewy body or TDP-43 pathologies.

Case RID 1393 was diagnosed with AD dementia antemortem, had a lower H:C, relatively atrophied hippocampi on MRI, thus, presented as *limbic-predominant AD*. At postmortem, case RID 1393 exhibited relatively high burdens of Aβ, tau, neuritic plaque and TDP-43 pathologies but not Lewy body pathology.

Case RID 1271 had AD dementia antemortem, lower BV:CSF (i.e., higher severity), widespread atrophy on MRI, thus, presented as *typical AD*. At postmortem, case RID 1271 demonstrated relatively high burdens of Aβ, tau and neuritic plaque pathologies, without evidence of Lewy body or TDP-43 pathologies.

Case RID 1425 was diagnosed with AD dementia antemortem, had higher BV:CSF (i.e., lower severity), relatively intact brain, thus, presented as *minimal-atrophy AD*. At postmortem, case RID 1425 had relatively high burdens of Aβ, tau and neuritic plaque pathologies, however, no Lewy body or TDP-43 pathologies.

### Antemortem atrophy subtypes and postmortem diagnosis

The primary neuropathological diagnosis was ADNC in all individuals. **Figure 4** shows that the secondary neuropathological diagnosis included none (n=8, 25.8%), amygdala Lewy bodies dementia (n=6, 19.3%), dementia with Lewy bodies (n=10, 32.2%), TDP-medial temporal lobe and hippocampal sclerosis (n=2, 6.4% for each), subdural hemorrhage, intracerebral hemorrhage, and SAL (n=1, 3.2% for each of the three diagnoses). Qualitatively, cases assigned to have TDP-43 in the medial temporal lobe or hippocampal sclerosis inclined towards limbic-predominant AD or typical AD. Cases assigned to have amygdala Lewy body pathology tended to be hippocampal-sparing AD or minimal-atrophy AD. The single isolated cases of intracerebral hemorrhage, subdural hemorrhage and SAL tended towards minimal-atrophy AD.

**Figure 4.**
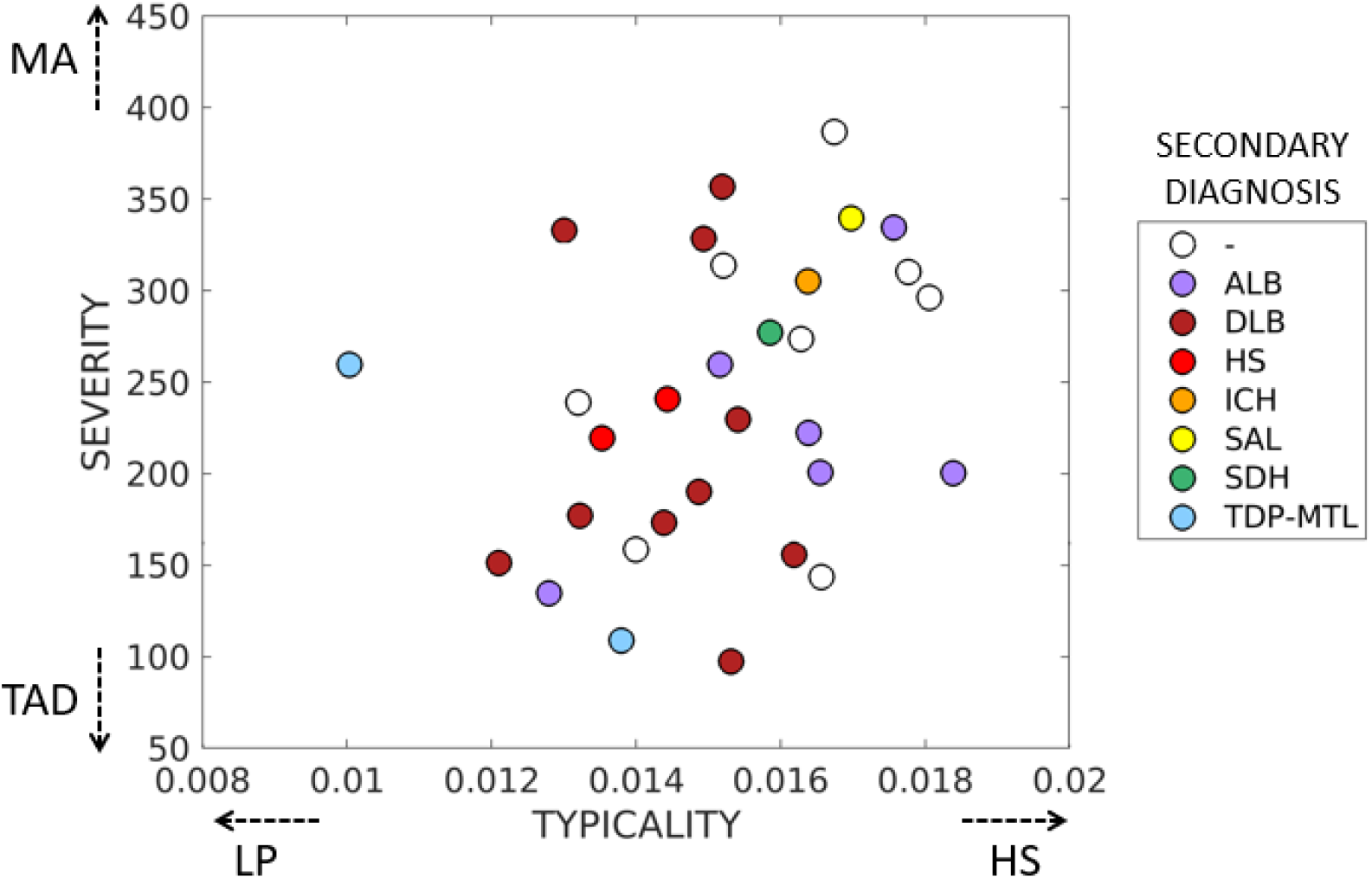
Secondary postmortem diagnosis superposed on antemortem MRI-based heterogeneity ALB=amygdala Lewy bodies; DLB=dementia with Lewy bodies; HS=hippocampal sclerosis; ICH=intracerebral hemorrhage; SDH=subdural hemorrhage; TDP-MTL=TAR DNA-binding protein in the medial temporal lobe; TAD=typical AD; HS=hippocampal-sparing AD; MA=minimal-atrophy AD; LP=limbic-predominant AD.

## Discussion

Our study investigated the relationship between antemortem atrophy subtypes and combinations of multiple postmortem (co)-pathologies in AD. Heterogeneity in AD is a multifaceted phenomenon involving combinations of protective factors, risk factors and concomitance of non-AD pathologies^7^. The relative contribution of (co)-pathologies to disease heterogeneity has been primarily reported from the postmortem (neuropathological) perspective^17,24–28^ with limited studies offering an antemortem (neuroimaging) perspective^29^. Our study serves as the first direct antemortem-to-postmortem validation, examining the interplay of these pathologies in atrophy subtypes of AD.

From the antemortem perspective, we treated biological heterogeneity in atrophy as a continuous phenomenon over the conventional approach of subgrouping individuals into subtypes^16^. For the first time, we examined an MRI-based operationalization of the conceptual framework for AD subtypes in terms of typicality and severity^7^, complementary to previous studies which categorizing individuals into distinct subtypes^30–33^. We observed a non-significant association between typicality and severity, suggesting that disease typicality (proxied by H:C) may not be influenced by disease staging or severity (proxied by BV:CSF). It is important, however, to note that our approach of treating typicality and severity separately may be rather simplistic. This is best exemplified by cases RID 1203 and RID 1452 (**Figure 2A**). Despite having a lower severity (higher BV:CSF), case RID 1203 was described as hippocampal-sparing AD rather than a minimal-atrophy AD. Thus, the combined contribution of typicality and severity must be factored in, i.e., every individual along the typicality dimension must also be treated in conjunction with the corresponding severity and vice-versa. Future studies should explore alternative operationalizations of this framework for comparison.

Our key finding was that antemortem typicality, but not severity, was associated with multiple forms of postmortem (co)-pathologies including tau, α-syn and TDP-43, both globally and regionally (**Table 2-3**). One reasoning for the lack of association between antemortem severity and postmortem (co)-pathologies could be that most individuals were at advanced disease stages (high AD neuropathologic change) with low variability in postmortem disease severity. Below we unravel the role of individual pathologies in relation to antemortem heterogeneity in atrophy.

### Amyloid pathology

We noted a global association between typicality and Thal Aβ stages, suggesting lower Aβ in hippocampal-sparing AD *atrophy* subtype, consistent with the recent meta-analysis evaluating the proportion of Aβ positivity in this subtype^7^. This result may be expected given that Aβ hallmark pathology in AD is rather diffuse, which may be indirectly associated with some degree of downstream atrophy^34^. Widespread Aβ accumulation is homogeneous with little regional specificity and may also explain why typicality was not associated with regional Aβ. This lack of regional associations likely reflects the lack of topographical correspondence between Aβ and atrophy, as evidence suggests a closer relationship between atrophy and tau than atrophy and Aβ^35–37^.

### Tau pathology

We did not observe a global association between typicality or severity and Braak tau stages, most likely due to little variability as all but two cases were at Braak stages V-VI. Regionally, however, limbic-predominant AD *atrophy* subtype was associated with higher tau NFT burden in the hippocampus. This is not surprising since tau NFT is an AD hallmark pathology affecting the hippocampus, particularly the dentate gyrus, which is known to contain the largest density of synapses^38^. Thus, higher tau burden may eventually be reflected in significant atrophy in the region, which is key characteristic of the limbic-predominant AD *atrophy* subtype. Conversely, hippocampal-sparing AD *atrophy* subtype was associated with lower tau NFT burden in the hippocampus. The association between atrophy and tau pathology in this subtype is not straightforward, owing to factors including the interval between assessments of these biomarkers^16^, regional non-specificity of atrophy and disagreement of subtyping methods based on these biomarkers^10^. Altogether, our study is useful in providing the direct link between antemortem atrophy and postmortem tau NFT, suggesting that hippocampal atrophy relative to neocortical atrophy can track postmortem tau NFT subtypes^17^.

### α-syn Lewy body pathology

We observed that limbic-predominant AD *atrophy* subtype and the limbic system in typical AD *atrophy* subtype may be more prone to α-syn Lewy body pathology. On one hand, this finding corroborates previous postmortem neuropathological studies showing increased Lewy body pathology in typical AD and limbic-predominant AD *tau* subtypes^17,24^. On the other hand, this finding contradicts other postmortem neuropathological studies showing increased Lewy body pathology in the hippocampal-sparing AD *tau* NFT subtype^25,26,29^. Two possible explanations arise for these diverging reports. Firstly, our regional analyses showed that the parahippocampal gyrus and superior/middle temporal gyri were both significantly associated with α-syn Lewy body pathology. While the former region could drive vulnerability of the limbic-predominant AD subtype to the pathology, the latter region may make hippocampal-sparing AD more susceptible. Secondly, most of the postmortem studies reporting presence of α-syn Lewy body pathology to date have characterized tau NFT subtypes, which are not necessarily and always interchangeable with atrophy subtypes in AD^10,16^. Therefore, deeper future *in vivo* investigations are warranted to confirm the role of α-syn Lewy body pathology in AD heterogeneity. Advanced aging, Braak staging (V-VI) and α-syn Lewy body staging (neocortical) in our cohort may support that the concomitance/interaction of tau and Lewy body pathologies may influence the atrophy observed in the limbic-predominant AD and typical AD atrophy subtypes since a lack of these factors has shown lack of limbic atrophy^39^.

### TDP-43 pathology

Our most robust findings included the association of limbic-predominant AD *atrophy* subtype to all forms of TDP-43 pathology globally and regionally. Limbic-predominant AD *tau* subtype has been described to be more prone to exhibiting TDP-43 in previous postmortem studies^17,24,26^. It is, thus, plausible for the limbic-predominant AD *atrophy* subtype to follow suit, given the topographical similarity between tau and atrophy patterns in limbic-predominant AD^16^. Congruent with the report from the recent meta-analysis^7^, our study provides the first antemortem-to-postmortem validation and evidence supporting the vulnerability of limbic-predominant AD *atrophy* subtype towards TDP-43. Globally, we observed a gradually increasing number of brain regions being affected by TDP-43 as one moves along the typicality dimension towards limbic-predominant AD. Regional examination revealed the strongest association between typicality and TDP-43 involving amygdala, which has been reported as an initial affected site by this pathology^28^, as well as other medial temporal lobe structures, as validated by a recent antemortem study^6^. As a common pathology affecting the hippocampus, TDP-43 is one if the main contributors of pathology, and the associated atrophy in the region may be detectable at least 10 years prior to death^40^. Given that individuals in our cohort were at advanced age (80 ± 6.7 y), it is also likely that some of them may have had Limbic-predominant age-related TDP-43 encephalopathy neuropathological changes (LATE-NC)^41^. In the absence of *in vivo* biomarkers assessing TDP-43, antemortem atrophy-based typicality (H:C) as a consistent correlate of postmortem TDP-43 in our study indicates the potential of this index as an antemortem proxy for the pathology.

Another main finding of our study was that both typicality and severity were regionally associated with concomitance of multiple (co)-pathologies. This relationship was such that hippocampal-sparing AD and minimal-atrophy AD were associated with higher concomitance of (co)-pathologies. There appears to be a region-specific effect, whereby some regions may accumulate greater number of pathologies while other regions may be spared. For example, hippocampal-sparing AD was associated with higher concomitance of (co)-pathologies particularly in several medial temporal structures, whereas minimal-atrophy AD was associated with higher pathological concomitance in the amygdala. At an individual-level, hippocampal structures including the dentate gyrus and CA1 notably demonstrated lower concomitance than other regions. The divergent reports on characteristic (co)-pathologies (e.g., to α-syn Lewy body pathology mentioned previously) associated with these two atrophy subtypes may be owing to the higher susceptibility of the subtypes to multiple or mixed pathologies. Higher concomitance, particularly in minimal-atrophy AD, raises the question of why this subtype shows minimal atrophy despite multiple pathologies. We speculate that one or more scenarios may explain this observation: (a) even though minimal-atrophy AD may accumulate multiple pathologies, the amount/burden of the pathologies may be relatively small, insufficient for overt atrophy to manifest; (b) even if the amount/burden of co-pathologies may be large, tau pathology (known to drive atrophy) may not have reached a high threshold; (c) co-occurrence of and/or interaction among multiple pathologies may affect an alternative biological process (e.g., metabolism, connectivity, etc.) in lieu of atrophy.

Finally, although qualitative, individual-level secondary postmortem diagnoses were informative. Two cases with lower H:C index (likely limbic-predominant AD) were assigned to have TDP-43 in the medial temporal region, consistent with our main quantitative findings. Two cases with lower H:C index (likely limbic-predominant AD) were assigned to have hippocampal sclerosis, which is known to correlate well with TDP-43 pathology^17,24^. Five out of six cases with relatively higher H:C index (likely hippocampal-sparing AD) were assigned to have amygdala Lewy body pathology. Whether/how the presence of this pathology plays a role in the disposition of hippocampal-sparing AD atrophy subtype to the pathology remains to be seen. Three cases with relatively higher H:C (likely hippocampal-sparing AD) and BV:CSF (likely minimal-atrophy AD) indices were assigned to have SAL, intracerebral hemorrhage and subdural hemorrhage, in line with our finding of higher pathological concomitance in these two subtypes. Overall, secondary diagnosis aided in providing greater confidence for some of our aforementioned findings.

Our study has some limitations. Firstly, the sample size of our cohort was limited, which may reduce the power to detect associations. However, our sample size was comparable to prior studies combining antemortem and postmortem data^8,9^, however, reduced the power to detect associations. Secondly, postmortem pathologies were only available as semi-quantitative scores (i.e., gross burden of pathology), which may not be as sensitive as quantitative scores obtained from digital histology techniques (e.g., specific counts, density or percentage of pathology per region). Finally, all data were sourced from the ADNI, known to have relatively strict inclusion criteria. Therefore, our current findings from our study would need to be further validated by future studies in the general population. Nevertheless, our study is strengthened by modeling subtypes as continuous phenomenon, avoiding introducing subtype-specific cutpoints^10,16^; and inclusion of group-as well as individual-level descriptions of the findings, lending itself to a deeper understanding of heterogeneity in AD.

In conclusion, we examined the relationship between antemortem atrophy subtypes and postmortem neuropathology in AD. Antemortem typicality on MRI shared a stronger global and regional association with individual and concomitance of postmortem (co)-pathologies including tau, α-synuclein and TDP-43, compared to antemortem severity. This suggests that the novel operationalization of typicality as a continuum is a promising proxy for spatial distribution of pathologies, irrespective of disease staging. Further, our findings show that it is critical to factor in contributions of core AD and non-AD (co)-pathologies for a deeper understanding of biological heterogeneity in AD, subsequently serving as an avenue for precision medicine and future multi-factorial therapies.

## Data Availability

Data used in this study are available from the Alzheimer's Disease Neuroimaging Initiative (ADNI) database (adni.loni.ucla.edu).

http://adni.loni.usc.edu/

## Acknowledgements

This study was funded by the Swedish Foundation for Strategic Research (SSF); the Strategic Research Programme in Neuroscience at Karolinska Institutet (StratNeuro); the Swedish Research Council (VR); the regional agreement on medical training and clinical research (ALF) between Stockholm County Council and Karolinska Institutet; Center for Innovative Medicine (CIMED); the Swedish Alzheimer Foundation; the Swedish Brain Foundation; the Åke Wiberg Foundation; Demensfonden; Stiftelsen Olle Engkvist Byggmästare; Birgitta och Sten Westerberg; Foundation for Geriatric Diseases at Karolinska Institutet; Loo och Hans Ostermans stiftelse för medicinsk forskning; Stiftelsen För Gamla Tjänarinnor; Gun & Bertil Stohnes Stiftelse. MJG is supported by the “Miguel Servet” program [CP19/00031] and a research grant [PI20/00613] of the Instituto de Salud Carlos III-Fondo Europeo de Desarrollo Regional (ISCIII-FEDER). The funding sources did not have any involvement in the study design; collection, analysis, and interpretation of data; writing of the report; and the decision to submit the article for publication.

Data collection and sharing for this study was funded by the Alzheimer’s Disease Neuroimaging Initiative (ADNI) (National Institutes of Health Grant U01 AG024904) and DOD ADNI (Department of Defense award number W81XWH-12-2-0012). ADNI is funded by the National Institute on Aging, the National Institute of Biomedical Imaging and Bioengineering, and through generous contributions from the following: Alzheimer’s Association; Alzheimer’s Drug Discovery Foundation; BioClinica, Inc.; Biogen Idec Inc.; Bristol-Myers Squibb Company; Eisai Inc.; Elan Pharmaceuticals, Inc.; Eli Lilly and Company; F. Hoffmann-La Roche Ltd and its affiliated company Genentech, Inc.; GE Healthcare; Innogenetics, N.V.; IXICO Ltd.; Janssen Alzheimer Immunotherapy Research & Development, LLC.; Johnson & Johnson Pharmaceutical Research & Development LLC.; Medpace, Inc.; Merck & Co., Inc.; Meso Scale Diagnostics, LLC.; NeuroRx Research; Novartis Pharmaceuticals Corporation; Pfizer Inc.; Piramal Imaging; Servier; Synarc Inc.; and Takeda Pharmaceutical Company. The Canadian Institutes of Health Research is providing funds to support ADNI clinical sites in Canada. Private sector contributions are facilitated by the Foundation for the National Institutes of Health (www.fnih.org). The grantee organization is the Northern California Institute for Research and Education, and the study is coordinated by the Alzheimer’s Disease Cooperative Study at the University of California, San Diego. ADNI data are disseminated by the Laboratory for Neuro Imaging at the University of California, Los Angeles.

Data used in this study were obtained from the Alzheimer’s Disease Neuroimaging Initiative (ADNI) database (adni.loni.ucla.edu). As such, the investigators within the ADNI contributed to the design and implementation of ADNI and/or provided data but did not participate in the analysis or writing of this report. A complete listing of ADNI investigators can be found at:http://adni.loni.ucla.edu/wp-content/uploads/how_to_apply/ADNI_Acknowledgement_List.pdf.

